# A follow-up study shows no new infections caused by patients with repeat positive of COVID-19 in Wuhan

**DOI:** 10.1101/2020.11.18.20232892

**Authors:** Xiaomin Wu, Zengmiao Wang, Zhenyu He, Yapin Li, Yating Wu, Huaiji Wang, Yonghong Liu, Fanghua Hao, Huaiyu Tian

**Affiliations:** Wuhan Center for Disease Control and Prevention, Wuhan, China; State Key Laboratory of Remote Sensing Science, Center for Global Change and Public Health, College of Global Change and Earth System Science, Beijing Normal University, Beijing, China; Central Theater Center for Disease Control and Prevention of PLA, Beijing, China; Central China Normal University, Wuhan, China

**Author notes:** Corresponding author. (Z.H.), (Z.W.). These authors contributed equally to this work (Xiaomin Wu, Zengmiao Wang, Zhenyu He, Yapin Li, Huaiyu Tian).

**Keywords:** COVID-19, Repeat positive, Close contact, Secondary infection

## Abstract

**Background:** It has been reported that a few recovered COVID-19 patients could suffer repeat positive, testing positive for the SARS-CoV-2 virus again after they were discharged from hospital. Understanding the epidemiological characteristics of patients with repeat positive is vital in preventing a second wave of COVID-19.

**Methods:** In this study, the epidemiological and clinical features for 20,280 COVID-19 patients from multiple centers between 31 December 2019 and 4 August 2020 in Wuhan were collected and followed. In addition, the RT-qPCR testing results for 4,079 individuals who had close contact with the patients suffering repeat positive were also obtained.

**Results:** 2,466 (12.16%) of 20,280 patients presented with a repeat positive of SARS-CoV-2 after they were discharged from hospital. 4,079 individuals had close contact with them. The PCR result were negative for the 4,079 individuals.

**Conclusions:** By a follow-up study in Wuhan, we show the basic characteristics of patients with repeat positive and no new infections caused by patients with repeat positive of COVID-19.

## Background

The ongoing severe acute respiratory syndrome-coronavirus 2 (SARS-CoV-2) pandemic has caused more than 49 million cases of coronavirus disease 2019 (COVID-19) illness and over 1.2 million people were dead duo to the coronavirus worldwide by November 2020(1). Guidelines from WHO on clinical management recommend that a clinically recovered COVID-19 patient should test negative for the virus twice, with testing supervised by trained professionals and conducted at least 24 hours apart, before being discharged from the hospital. It has been reported that a few recovered COVID-19 patients could suffer repeat positive, testing positive for the virus again within the 14-day isolation period(2–4). The number of patients reported to have had repeat positive of COVID-19 is small and the duration of follow-up has been short. Understanding the epidemiological characteristics of patients with repeat positive is vital in preventing a second wave of COVID-19.

## Methods

### Study description

The COVID-19 outbreak was first reported in Wuhan(5) and had lasted more than 3 months by 5 April 2020, after which date there were no further locally acquired infections. During the outbreak in Wuhan, there were a total of 50333 confirmed COVID-19 cases with 3869 deaths, and 46464 patients have been clinically cured and discharged according to WHO guidelines(6). After being discharged from hospital, patients in Wuhan continue to be isolated in rehabilitation center for 14 days and at home for another 14 days to prevent reinfection, followed up and then regularly revisited at hospitals. Here we collect all the information on SARS-CoV-2 patients with repeat positive between 31 December 2019 and 4 August 2020 across 84 hospitals in Wuhan, presenting epidemiological and clinical features.

### Data collection

Cases of COVID-19 was diagnosed and the illness severity was defined according to the Chinese management guideline for COVID-19 (the sixth edition) published by National Health Commission of the People’s Republic of China. All first diagnosis cases of COVID-19 were confirmed according to positive respiratory RT-PCR tests. Repeat positive were confirmed by digestive (anal swab) and respiratory positive RT-PCR tests. Samples by Nasopharyngeal swab or Pharynx swab were collected and tested for SARS-CoV-2, following WHO guidelines.

The discharge criteria of the recovered patients included: body temperature is back to normal for more than three days, respiratory symptoms improve obviously, pulmonary imaging shows obvious absorption of inflammation, and nucleic acid tests negative for respiratory tract pathogen twice consecutively (sampling interval being at least 24 hours). Those who meet the above criteria can be discharged. After discharge, it is recommended for the patients to continue 14 days of isolation management and health monitoring, wear a mask, live in a single room with good ventilation. The patients are required to return to the hospitals for follow-up and revisit in two and four weeks after discharge.

### Statistical analysis

In this study, categorical variables were presented as numbers and percentages, and continuous variables were presented using median (interquartile range, IQR). Student *t*-test was used to compare continuous variables in two groups. Multivariate logistic regression model was used to determine the associated factors. All statistical analyses were performed using R version 3.4.1. *P* value less than 0.05 was considered statistically significant.

## Results

In total, 20,280 patients were collected and followed. Among them, 2,466(12.16%) patients presented with a repeat positive of SARS-CoV-2 after they were discharged from hospital. The demographic and epidemiological characteristics of the these patients are similar to the first infections (shown in Table 1). The median treatment time for patients without repeat positive in hospital was 15.71days (interquartile range [IQR], 9.83 to 23.21) and the median treatment time for the first infection in patients with a repeat positive was 18.54 days (interquartile range [IQR], 11.96 to 27.04). The treatment time for first infection in patients with a repeat positive was significantly longer than that in patients without a repeat positive (p-value <0.001from Student *t*-test). For the repeat positive, the median treatment time was 10.63 days (interquartile range [IQR], 7.67 to 15.63). The treatment time for the repeat positive was shorter than that for the first infection among the patients with a repeat positive. The median time from discharge from hospital to the start of a second positive was 11.00 days (IQR, 9.00 to 17.00). A total of 56.12% were female. The median age of the patients was 56.00 years (IQR, 42.25 to 65.00). In addition, more than half of the patients (50.9%, n=1256) were aged between 50 and 70 years (accounted for 15.6% of the total population in Wuhan). Symptoms of first infection for patients with repeat positive were 0.24% asymptomatic, 49.67% mild, 35.59% moderate, 12.05% severe and 2.40% critical. For the symptoms of the second positive, of 2,466, 193(7.83%) patients had fever, cough or shortness of breath. Specifically, 158(6.41%) patients had fever, 59 (2.39%) patients had cough and 18(0.73%) patients had shortness of breath. 32(1.30%) patients had both fever and cough. 1(0.04%) patient had both cough and shortness of breath. 9 (0.36%) patients had both fever and shortness of breath. No patients had three types of symptom at the same time. 11 of 2,466 patients were passed away.

**Table 1.**
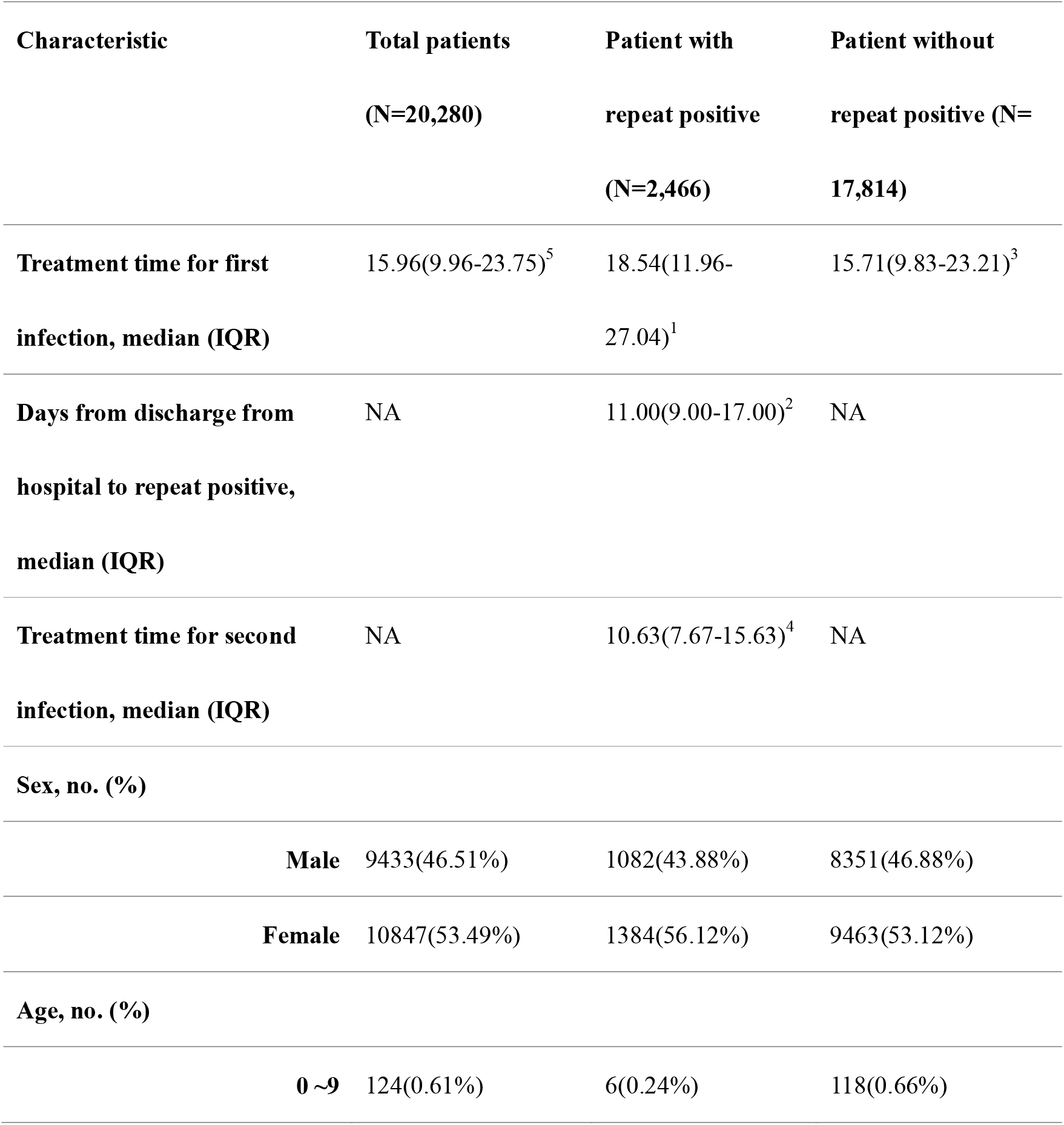
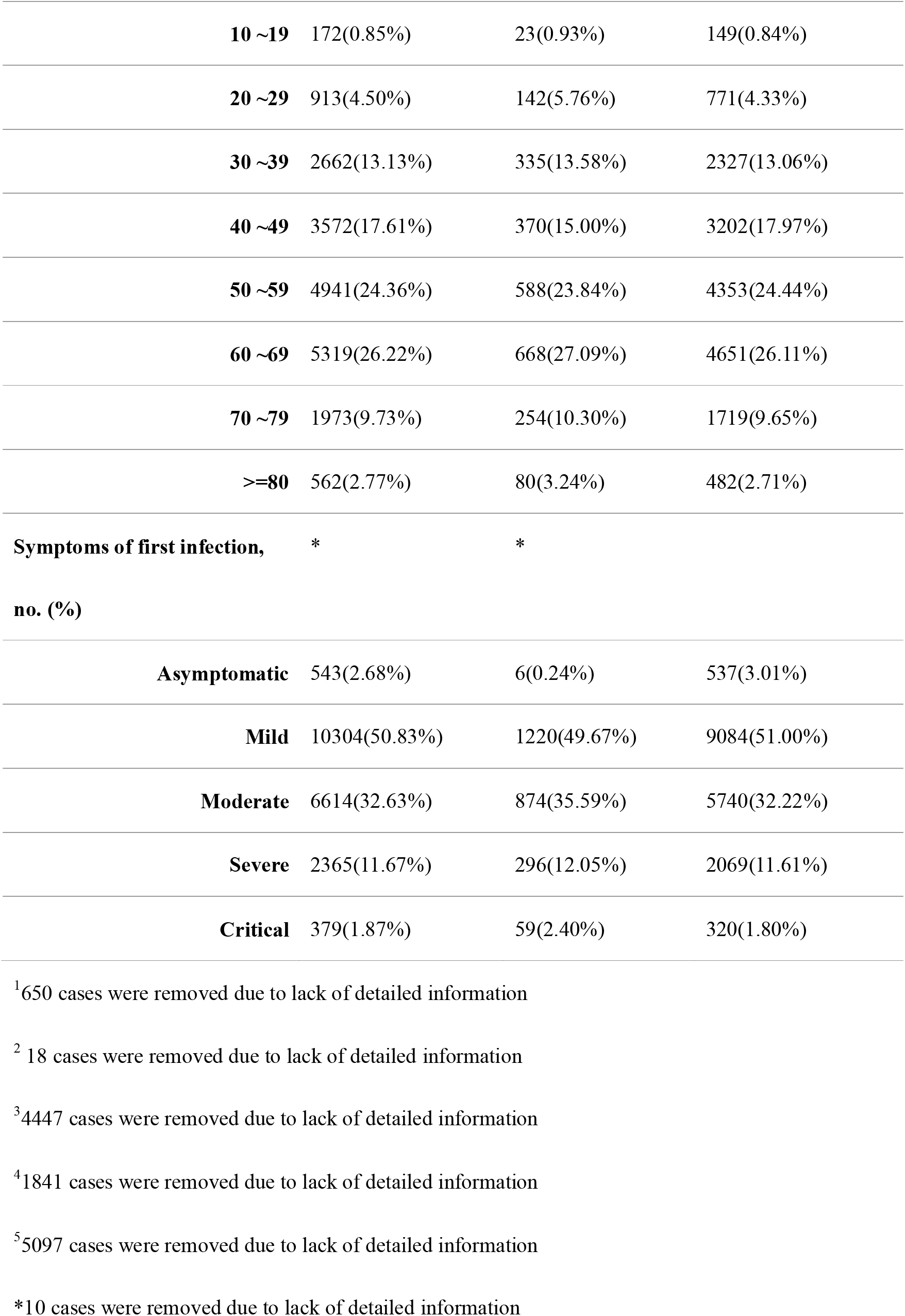
Characteristics of SARS-CoV-2 positive cases by presence of repeat positive, Wuhan, China, December 2019-August 2020.

A logistic regression model was used to check the factors related to a repeat positive. The age had no effects (coeff.=0.001, *P*=0.379) on the probability of a repeat positive. Interestingly, we found that the gender had a significantly role (coeff.=-0.119, *P*=0.005). The males had a lower risk of developing a repeat positive compared to females.

To investigate whether the patients with repeat positive can cause the new infections, we collected the PCR results for the people who had close contact with patients with repeat positive. Of 2,466 patients with repeat positive, 1,201 patients were tested positive in rehabilitation center and there were no individuals who had close contact with them. So these 1,201 patients had no chance to infect healthy persons. 1,265 patients were tested positive after they went back home. 4,079 individuals had close contact with them. The PCR result were negative for the 4,079 individuals. This indicates that the patients with repeat positive don’t cause new infections.

## Discussion

Although several studies have reported patients with repeat positive of COVID-19, most are based on a limited sample and the proportion of SARS-CoV-2 reactivation ranged from 9% (5/55)(2) to 14.5% (38/262)(4). This study retrospectively analyzed clinical data in a cohort of 20,280 patients in Wuhan. We confirmed that in 12.16% of COVID-19 patients, SARS-CoV-2 could be isolated again after discharge from hospital. The time from SARS-CoV-2 negative to positive ranged from 1 to 165 days, suggesting that recovered patients still may be virus carriers and require additional round of viral detection and isolation. Note that the repeat positive could also be the virus fragment coming from the first infection because the PCR test only detects the fragments of SARS-CoV-2 genome, not viable virus. Another study shown that no infectious strain could be obtained by culture and no full-length viral genomes could be sequenced from 87 re-positive cases(7). These may explain why there were no secondary infection. Although this study does not completely exclude the possibility of reinfection, however, given the 28-day isolation and 84.07% (2058/2448) of the repeat positive occurred during this isolation in Wuhan, most of these patients have apparently not caused new infections after discharge from hospital in Wuhan.

## Conclusions

In this follow-up study in Wuhan, we show the basic characteristics of patients with repeat positive and no new infections caused by patients with repeat positive of COVID-19.

## Data Availability

The data supporting the findings of this study are available within the article.

## Term definition

Patient with repeat positive: the patient was test positive for SARS-CoV-2 again after being discharged from hospital.
Treatment time: time interval of a patient being hospitalized.
Close contact: the individuals living in the same houses with the patients.

## Ethics approval and consent to participate

The epidemiological and clinical data collection was exempt from Institutional Review Boards, because it was part of public health investigation for COVID-19, issued by the National Health Commission of the People’s Republic of China (http://www.nhc.gov.cn/jkj/s3577/202003/4856d5b0458141fa9f376853224d41d7.shtml). This study was approved by the Institutional Review Boards from Wuhan Center for Disease Control and Prevention.

## Consent for publication

Not applicable.

## Availability of data and materials

The data supporting the findings of this study are available within the article.

## Author contributions

X.W., F.H., Y.L., H.T, Z.H., H.W. designed the study. X.W., Y.W., Z.H. collected the statistical data. Y.L. and Z.W conducted the analyses. H.T. wrote the manuscript. All authors read and approved the manuscript.

## Acknowledgements

We thank the many thousands of CDC staff and local health workers in China who collected data.

